# WHY SOME HESITATE MORE: CROSS-CULTURAL VARIATION IN CONSPIRACY BELIEFS, BELIEF IN SCIENCE, AND VACCINE ATTITUDES

**DOI:** 10.1101/2021.07.09.21260228

**Authors:** Gul Deniz Salali, Mete Sefa Uysal

## Abstract

**Background:** Countries differ in their levels of vaccine hesitancy (a delay in acceptance or refusal of vaccines), trust in vaccines, and acceptance of new vaccines. In this paper, we examine the factors contributing to the cross-cultural variation in vaccine attitudes, measured by levels of 1) general vaccine hesitancy, 2) trust in vaccines, and 3) COVID-19 vaccine acceptance.

**Methods:** We examined the relative effect of conspiracy mentality, belief in COVID-19 conspiracies, and belief in science on the above-mentioned vaccine attitudes in the UK (n= 1533), US (n= 1550), and Turkey (n= 1567) through a quota-sampled online survey to match the population for age, gender, ethnicity, and education level.

**Results:** We found that belief in COVID-19 conspiracies and conspiracy mentality were the strongest predictors of general vaccine hesitancy across all three countries. Belief in science had the largest positive effect on general vaccine trust and COVID-19 vaccine acceptance. Although participants in Turkey demonstrated the lowest level of vaccine trust, their belief in science score was significantly higher than participants in the US, suggesting that belief in science cannot explain the cross-cultural variation in vaccine trust. The mean levels of conspiracy mentality and agreement with COVID-19 conspiracies were consistent with the country-level differences in general and COVID-19 vaccine attitudes. Demographic variables did not predict vaccine attitudes as much as belief in conspiracies and science.

**Conclusions:** Our findings suggest that cross-cultural variation in vaccine hesitancy, vaccine trust, and COVID-19 vaccine acceptance rates are mainly driven by differences in the prevalence of conspiratorial thinking across countries.

## Introduction

Why do we observe variation in vaccine hesitancy, vaccine trust, and acceptance of new vaccines across countries? People in some countries are more hesitant about getting vaccinated than people in other countries (de Figueiredo, Simas, Karafillakis, Paterson, & Larson, 2020). Understanding such variation has become increasingly important in the face of the ongoing COVID-19 pandemic. A sufficient response to the pandemic requires not only individual countries successfully vaccinating their populations, but all countries across the world having successful vaccination programs. Besides the vaccine supply problem, vaccine hesitancy is the largest obstacle to achieving herd immunity in many countries. Cross-cultural analyses of COVID-19 vaccine acceptance rates indicate that acceptance rates vary from 24% to over 90% across the world (Lazarus et al., 2021; Sallam, 2021). The estimates suggest that with a 100% efficient vaccine, 60-72% of people need to be vaccinated against COVID-19 to achieve herd immunity, depending on the population-specific infection rates (Anderson, Vegvari, Truscott, & Collyer, 2020; Kwok, Lai, Wei, Wong, & Tang, 2020). Therefore, it is important to focus in particular on countries where reported vaccine hesitancy and COVID-19 infection rates are high and to ask what psychological and cultural factors contribute to the cross-cultural variation in vaccine acceptance. In this paper, we address this question and examine the factors affecting general vaccine hesitancy and trust and COVID-19 vaccine acceptance in Turkey, the UK, and the US.

Previous research has identified several psychological factors contributing to vaccine attitudes. Among these, conspiracy beliefs have been found to be strongly associated with anti-vaccine views and negative attitudes towards vaccines (Bertin, Nera, & Delouvée, 2020; Hornsey, Harris, & Fielding, 2018; Jolley & Douglas, 2014; Lewandowsky, Gignac, & Oberauer, 2013). Conspiracy beliefs find explanation for events happening in the world in the supposed unlawful or harmful acts of a secret, powerful group of people (Zonis & Joseph, 1994). Recent studies have also found that acceptance of COVID-19 vaccines is influenced by susceptibility to misinformation and COVID-19 conspiracy beliefs (Freeman et al., 2021; Roozenbeek et al., 2020; Salali & Uysal, 2020; Sallam et al., 2021). There is variation in the level of belief in conspiracies across countries (Bruder, Haffke, Neave, Nouripanah, & Imhoff, 2013). Conspiratorial thinking, for example, is relatively high in Middle Eastern countries (Zonis & Joseph, 1994). Although conspiratorial thinking is linked to vaccine hesitancy, it is unclear to date whether cross-cultural variation in vaccine hesitancy is driven by the observed variation in conspiratorial thinking.

Previous research has found that if a person believes in one conspiracy, he or she is more likely to believe in others (Goertzel, 1994). Therefore, some people are considered to be more susceptible to conspiracy beliefs and have a general *conspiracy mentality* (Bruder et al., 2013). Since the start of the COVID-19 pandemic, a myriad of conspiracy theories have emerged (Freeman et al., 2020). Although we predict people with high conspiracy mentality scores to believe in individual conspiracy theories specific to COVID-19 more than others, it is possible that a general conspiratorial worldview influences vaccination attitudes differently than specific conspiracy beliefs about the origin of COVID-19. For example, a person may believe that many important things happen in the world without the knowledge of the public, but the influence of this belief on their vaccination decisions may be lower than the influence of a more specific conspiracy belief, such as a belief that the coronavirus was manufactured to reduce the world population. Therefore, one aim of this study is to investigate and compare the relative effects of a general conspiracy mentality and specific conspiracy beliefs regarding COVID-19 on vaccine attitudes.

An additional factor that has been demonstrated to affect vaccine attitudes is beliefs about science and scientists. Countries with a stronger consensus in trust in science and scientists demonstrate an overall higher confidence in vaccines (Sturgis, Brunton-Smith, & Jackson, 2021). Greater trust in scientists is also associated with lower susceptibility to COVID-19 misinformation, which is linked to vaccine acceptance (Roozenbeek et al., 2020). Nevertheless, there is a lack of information on the relative importance of conspiracy beliefs and belief in science on vaccine attitudes across countries. It is important to stress here that the factors related to vaccine hesitancy may differ from those that contribute to trust in vaccines and acceptance of new vaccines. Although most studies have examined either the reasons for vaccine hesitancy or trust in vaccines, we believe it is important to examine the effects of several factors on vaccine hesitancy, vaccine trust, and acceptance of new vaccines separately. For example, while people’s perceptions of science and scientists may strongly correlate with their trust in the effectiveness of vaccines, such perceptions may be irrelevant to their reasons for vaccine hesitancy. Moreover, the factors influencing attitudes towards new vaccines, such as COVID-19 vaccines, may differ from those related to attitudes towards vaccines in general. Therefore, in this study, we aim to separately examine the relative effects of conspiracy mentality, COVID-19 conspiracy beliefs, and belief in science on i) general vaccine hesitancy (level of agreement with statements related to unforeseen vaccine side effects, preference for natural immunity, or commercial profits), ii) vaccine trust (agreement with the safety and effectiveness of vaccines), and iii) COVID-19 vaccine acceptance.

Although a few previous studies have found high cross-cultural variation in vaccine attitudes (de Figueiredo et al., 2020; Gallup, 2019), only one study has comprehensively investigated the psychological roots of this variation (Hornsey et al., 2018). Moreover, the reasons for vaccine hesitancy in low-and middle-income countries are largely understudied (Bono et al., 2021; Simas & Larson, 2021). It is important to understand the country-level psychological factors contributing to the variation in vaccine attitudes in an ever-connected world. Given the ongoing success of the COVID-19 vaccination program in the UK, we predict attitudes towards vaccination in the UK to be more positive than those in the US. The difference in vaccine attitudes between these two countries may be driven by differences in levels of conspiratorial thinking. Our previous research in May 2020 found COVID-19 vaccine acceptance rates to be much lower in Turkey than in the UK (Salali & Uysal, 2020). According to our results, the probability of acceptance increased if a person believed in the natural origin of the virus, as opposed to a man-made origin, a belief that many more participants in Turkey held than in the UK (Salali & Uysal, 2020). Conspiracy beliefs are a widespread feature of culture and politics in Turkey, and conspiracy mentality is more prevalent among Turkish people than people in Western Europe and the US (Bruder et al., 2013). However, it remains unclear how conspiracy mentality affects vaccination decisions in this country.

Taken together, we aim to understand the following:

1. Cross-cultural variation in vaccine hesitancy, trust in vaccines, and COVID-19 vaccine acceptance: Based on previous findings, we predict vaccine hesitancy to be highest in Turkey and trust in vaccines and COVID-19 vaccine uptake to be highest in the UK.
2. The relative importance of conspiracy mentality, COVID-19 conspiracy beliefs, and belief in science in explaining the variation in vaccine attitudes: We predict that the observed cross-cultural variation in vaccine hesitancy is mainly driven by the level of conspiratorial thinking in the three countries. Variation in belief in science may explain cross-cultural variation in vaccine trust and acceptance of new vaccines, but not necessarily vaccine hesitancy.
3. The relative effect of several demographic variables (age, gender, political orientation, education level, ethnicity—for the UK and US) in each country on vaccine attitudes.

## Methods

### Study sample

We conducted an anonymous online survey with a quota sample of participants in the UK (n= 1533), US (n= 1550), and Turkey (n= 1567) from March 9^th^ to April 8^th^, 2021. The quotas were based on population estimate data for age, gender, and education level for each country (and ethnicity for the UK and the US), and participants were recruited by Qualtrics. We excluded participants who i) did not reply “yes” to the initial check question on whether they have spent most of their childhood in the country in which they were participating or ii) did not pass the attention check questions during the survey. Table 1 lists all of the demographic variables and their mean values for each country. Informed consent was collected from all participants. The study was approved by the UCL Research Ethics Committee (ID: 13121/003).

**Table 1.** Demographic variables

|  | Mean (SD)/n(%)<br>UK | Mean<br>(SD)/n(%)<br>US | Mean (SD)/n(%)<br>Turkey |
| --- | --- | --- | --- |
| *Age (years)* | 45.13 (16.16) | 44.69 (16.75) | 40.73 (14.94) |
| 18-24 | 11.5% | 14.0% | 15.1% |
| 25-34 | 19.6% | 19.0% | 24.1% |
| 35-44 | 18.4% | 18.3% | 22.9% |
| 45-54 | 19.6% | 16.7% | 17.4% |
| 55-64 | 16.8% | 17.2% | 13.2% |
| 65+ | 14.2% | 14.6% | 7.3% |
| *Gender* |  |  |  |
| Female | 792 (51.7%) | 799 (51.5%) | 777 (49.6%) |
| Male | 732 (47.7%) | 736 (47.5%) | 771 (49.2%) |
| Non-Binary | 8 (0.5%) | 10 (0.6%) | 18 (1.1%) |
| Prefer not to say | 1 (0.1%) | 5 (0.3%) | 1 (0.1%) |
| *Education* |  |  |  |
| Below Graduate | 708 (46.1%) | 1011 (65.2%) | 1180 (75.3%) |
| Graduate Degree | 586 (38.2%) | 341 (22.0%) | 342 (21.8%) |
| Postgraduate Degree | 239 (15.6%) | 198 (12.8%) | 45 (2.9%) |
| *Political Orientation (0 = Left, 10 = Right)* | 4.89 (2.09) | 5.30 (2.82) | 5.24 (2.88) |
| *Financial Satisfaction (0-10)* | 5.43 (2.57) | 5.09 (2.95) | 4.90 (2.84) |
| *Ethnicity* |  |  |  |
| White British | 1333 (87%) |  |  |
| Black/ African / Caribbean | 43 (3%) |  |  |
| Asian | 108 (7%) |  |  |
| Other Ethnic Groups | 49 (3%) |  |  |
| Non-Hispanic White |  | 932 (60%) |  |
| Hispanic / Latino |  | 249 (16%) |  |
| Black / African American |  | 208 (13%) |  |
| Asian |  | 92 (6%) |  |
| Other |  | 69 (4%) |  |

### Psychological variables

Table 2 presents the psychological variables that were measured in the survey, the statements employed to capture the variables, the response scales, Cronbach’s alphas, and the country-level means or percentages. All survey questions were translated into Turkish by the authors and sent to native speakers for an additional check.

**Table 2.**
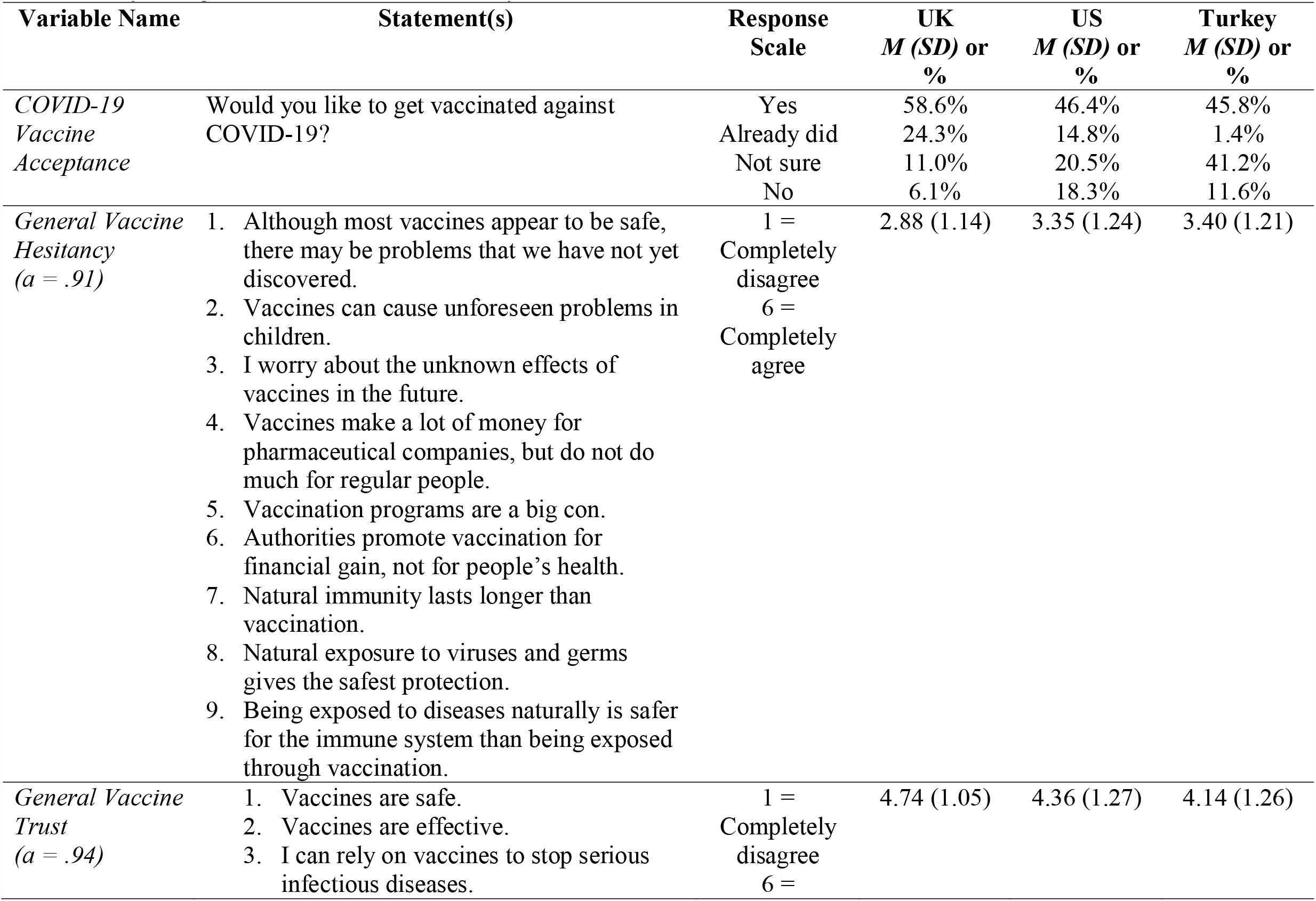
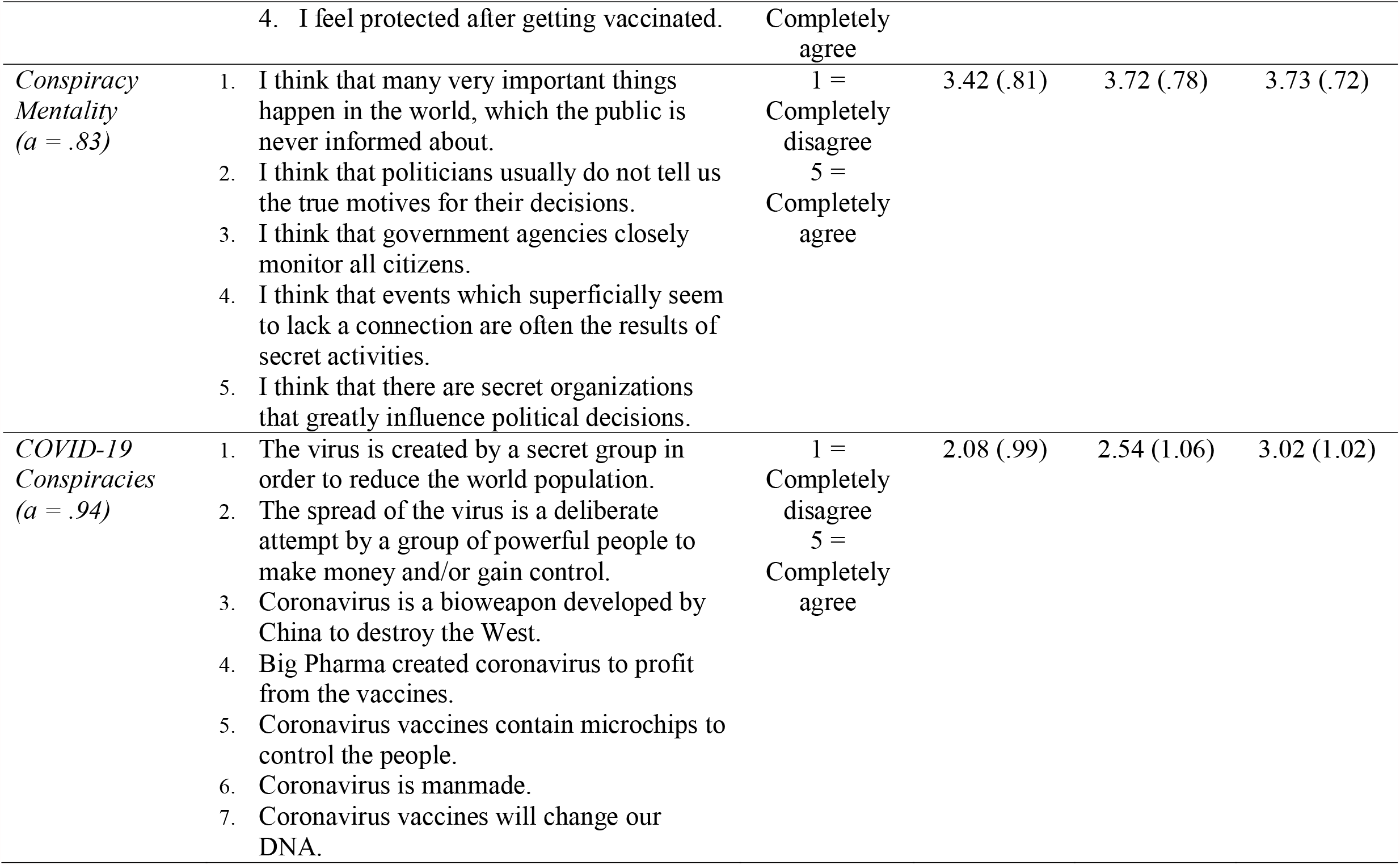
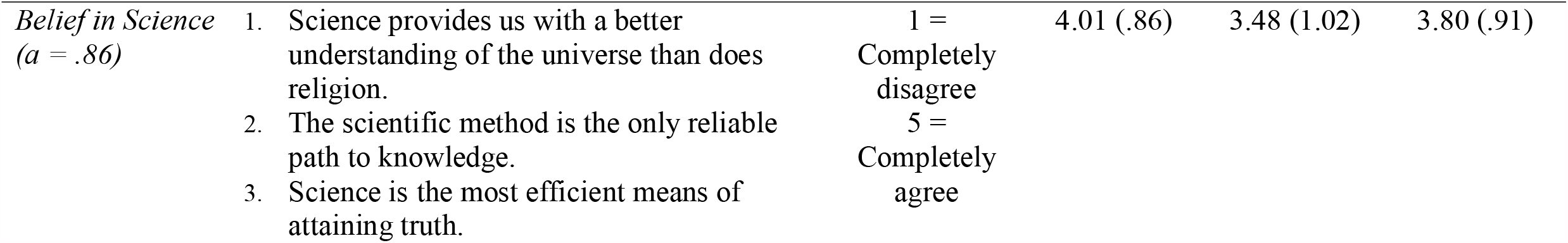
Psychological variables used in the study

| Variable Name | Statement(s) | Response Scale | UK<br>*M (SD)* or<br>% | US<br>*M (SD)* or<br>% | Turkey<br>*M (SD)* or<br>% |
| --- | --- | --- | --- | --- | --- |
| *COVID-19 Vaccine Acceptance* | Would you like to get vaccinated against COVID-19? | Yes<br>Already did<br>Not sure<br>No | 58.6%<br>24.3%<br>11.0%<br>6.1% | 46.4%<br>14.8%<br>20.5%<br>18.3% | 45.8%<br>1.4%<br>41.2%<br>11.6% |
| *General Vaccine Hesitancy*<br>(*a* = .91) | 1. Although most vaccines appear to be safe, there may be problems that we have not yet discovered.<br>2. Vaccines can cause unforeseen problems in children.<br>3. I worry about the unknown effects of vaccines in the future.<br>4. Vaccines make a lot of money for pharmaceutical companies, but do not do much for regular people.<br>5. Vaccination programs are a big con.<br>6. Authorities promote vaccination for financial gain, not for people's health.<br>7. Natural immunity lasts longer than vaccination.<br>8. Natural exposure to viruses and germs gives the safest protection.<br>9. Being exposed to diseases naturally is safer for the immune system than being exposed through vaccination. | 1 = Completely disagree<br>6 = Completely agree | 2.88 (1.14) | 3.35 (1.24) | 3.40 (1.21) |
| *General Vaccine Trust*<br>(*a* = .94) | 1. Vaccines are safe.<br>2. Vaccines are effective.<br>3. I can rely on vaccines to stop serious infectious diseases. | 1 = Completely disagree<br>6 = | 4.74 (1.05) | 4.36 (1.27) | 4.14 (1.26) |
|  | 4. I feel protected after getting vaccinated. | Completely agree |  |  |  |
| *Conspiracy Mentality*<br>($a = .83$) | 1. I think that many very important things happen in the world, which the public is never informed about. | 1 = Completely disagree | 3.42 (.81) | 3.72 (.78) | 3.73 (.72) |
|  | 2. I think that politicians usually do not tell us the true motives for their decisions. | 5 = Completely agree |  |  |  |
|  | 3. I think that government agencies closely monitor all citizens. |  |  |  |  |
|  | 4. I think that events which superficially seem to lack a connection are often the results of secret activities. |  |  |  |  |
|  | 5. I think that there are secret organizations that greatly influence political decisions. |  |  |  |  |
| *COVID-19 Conspiracies*<br>($a = .94$) | 1. The virus is created by a secret group in order to reduce the world population. | 1 = Completely disagree | 2.08 (.99) | 2.54 (1.06) | 3.02 (1.02) |
|  | 2. The spread of the virus is a deliberate attempt by a group of powerful people to make money and/or gain control. | 5 = Completely agree |  |  |  |
|  | 3. Coronavirus is a bioweapon developed by China to destroy the West. |  |  |  |  |
|  | 4. Big Pharma created coronavirus to profit from the vaccines. |  |  |  |  |
|  | 5. Coronavirus vaccines contain microchips to control the people. |  |  |  |  |
|  | 6. Coronavirus is manmade. |  |  |  |  |
|  | 7. Coronavirus vaccines will change our DNA. |  |  |  |  |
| *Belief in Science*<br>($\alpha = .86$) | 1. Science provides us with a better understanding of the universe than does religion. | 1 = Completely disagree | 4.01 (.86) | 3.48 (1.02) | 3.80 (.91) |
|  | 2. The scientific method is the only reliable path to knowledge. | 5 = Completely agree |  |  |  |
|  | 3. Science is the most efficient means of attaining truth. |  |  |  |  |

#### COVID-19 vaccine acceptance

We measured whether the participants want to receive a COVID-19 vaccine.

#### General vaccine hesitancy

We measured vaccine hesitancy using the “worries about unforeseen future effects”, “concerns about commercial profiteering”, and “preference for natural immunity” subscales of the VAX scale (Martin & Petrie, 2017).

#### General vaccine trust

We measured vaccine trust by asking participants about their agreement with positive statements concerning the safety and effectiveness of the vaccines, using items from the “trust of vaccine benefit” subscale of the VAX scale (Martin & Petrie, 2017).

#### Conspiracy mentality

We used a general conspiracy mentality scale (Bruder et al., 2013). We chose this scale as it is designed to measure the general tendency to believe in conspiracies, regardless of cultural factors associated with more specific conspiracy statements.

#### COVID-19-specific conspiracy beliefs

We chose, among the COVID-19 conspiracy beliefs (Freeman et al., 2020), conspiracy statements that are most popular (according to their appearance on the news, social media, and WhatsApp groups) across the three countries at the time of the study.

#### Belief in science

We measured belief in science using a selection of items from the Belief in Science Scale based on their factor loadings (Farias, Newheiser, Kahane, & de Toledo, 2013).

### Statistical Analysis

We first calculated bivariate relationships across all variables for each country. We then conducted multiple linear regression analyses to examine the predictive power of each of the psychological and demographic variables on general vaccine hesitancy and vaccine trust in each country. We calculated the effect of each variable on the probability of COVID-19 vaccine acceptance (willing to be vaccinated/already vaccinated coded as 1, not sure/no coded as 0) by logistic regression analysis. We used ANOVA followed by Tukey’s honestly significant difference (HSD) tests to compare the mean levels of general vaccine hesitancy, vaccine trust, COVID-19 vaccine acceptance, conspiracy mentality, belief in COVID-19 conspiracies, and belief in science among the three countries. We conducted the analyses in R statistical software.

## Results

### General and COVID-19 vaccine attitudes vary across countries

Participants in the UK had a significantly lower mean vaccine hesitancy score than the participants in the US and Turkey (Table 2, Tukey’s HSD, p< 0.001). There was no difference in the mean vaccine hesitancy scores between the US and Turkey (Table 2, Tukey’s HSD, p= 0.5). General vaccine trust levels were highest in the UK and lowest in Turkey (Table 2, Tukey’s HSD: p< 0.001 for UK-US, UK-Turkey and US-Turkey). Likewise, COVID-19 vaccine acceptance rates were highest in the UK and lowest in Turkey (Table 2, Tukey’s HSD: p< 0.001 for UK-US, UK-Turkey and US-Turkey). The proportion of participants who refused to get vaccinated against COVID-19 was highest in the US (Table 2).

### The strongest predictor of general vaccine hesitancy across the three countries is belief in COVID-19 conspiracies

There was a strong positive correlation between conspiracy mentality, belief in COVID-19 conspiracies, and general vaccine hesitancy across the three countries (Figure 1a-b, Table 3a-c). Controlling for other factors, belief in COVID-19 conspiracies had the largest effect on general vaccine hesitancy (Figure 1a, Table 4a-c). Although its effect size was smaller compared to belief in COVID-19 conspiracies, conspiracy mentality had the second largest effect on general vaccine hesitancy across all three countries (Figure 1b, Table 4a-c).

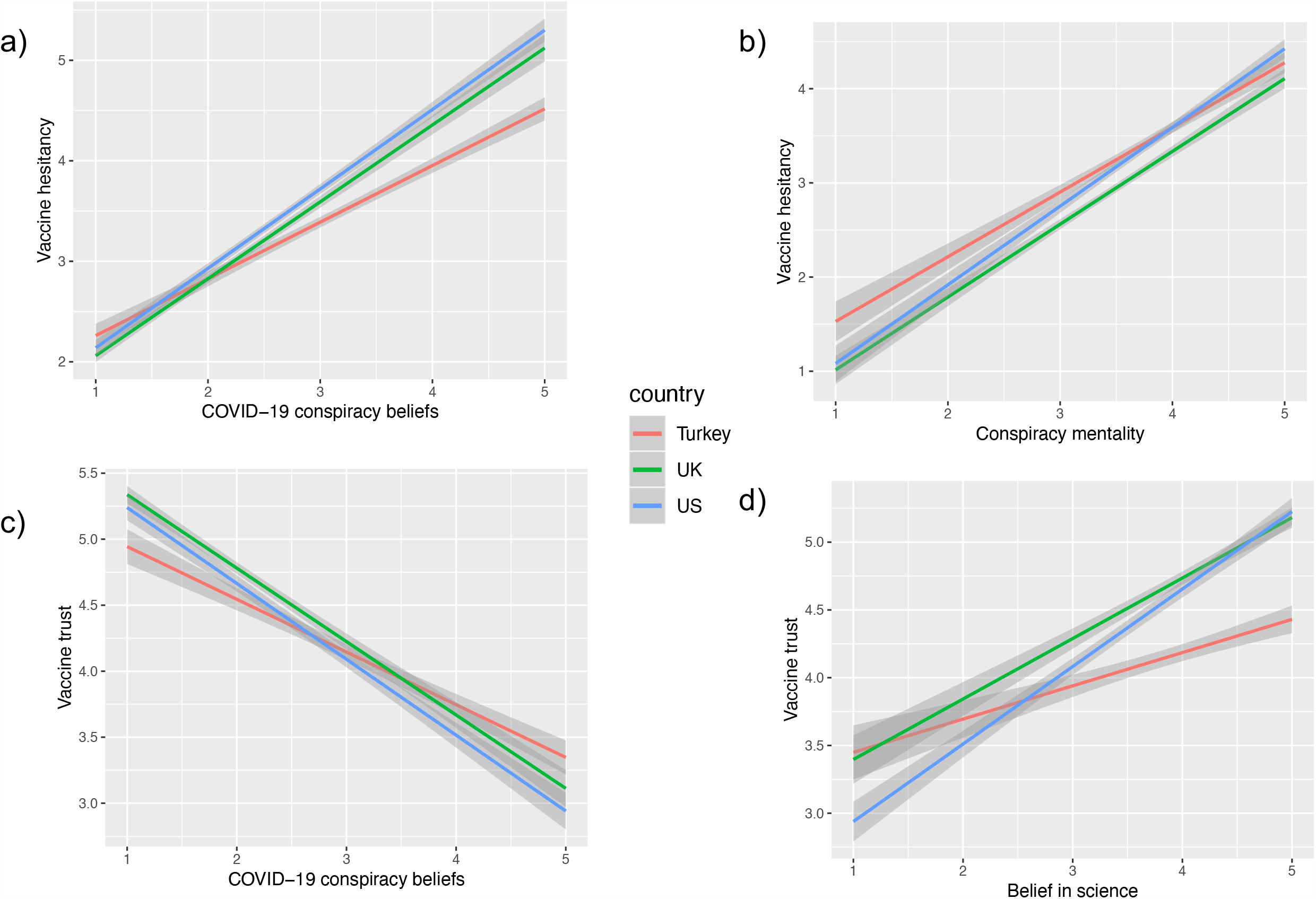

**Table 3a.** Bivariate correlations between variables (UK)

|  | 1 | 2 | 3 | 4 | 5 | 6 | 7 | 8 |
| --- | --- | --- | --- | --- | --- | --- | --- | --- |
| 1. COVID-19 Vaccine Acceptance | - | -.53*** | .59*** | -.28*** | -.37*** | .26*** | -.03 | .17*** |
| 2. General Vaccine Hesitancy |  | - | -.64*** | .55*** | .66*** | -.28*** | .20*** | -.14*** |
| 3. General Vaccine Trust |  |  | - | -.34*** | -.53*** | .37*** | -.02 | .21*** |
| 4. Conspiracy Mentality |  |  |  | - | .53*** | -.11*** | .04 | -.19*** |
| 5. COVID-19 Conspiracies |  |  |  |  | - | -.29*** | .26*** | -.08** |
| 6. Belief in Science |  |  |  |  |  | - | -.08** | .07** |
| 7. Political Orientation |  |  |  |  |  |  | - | .20*** |
| 8. Financial Satisfaction |  |  |  |  |  |  |  | - |

Note. \*\*\* $p < .001$ , \*\* $p < .01$ ; Political orientation: 1 = left, 10 = right

**Table 3b.** Bivariate correlations between variables (US)

|  | 1 | 2 | 3 | 4 | 5 | 6 | 7 | 8 |
| --- | --- | --- | --- | --- | --- | --- | --- | --- |
| 1. COVID-19 Vaccine Acceptance | - | -.51*** | .64*** | -.29*** | -.41*** | .37*** | -.09** | .20*** |
| 2. General Vaccine Hesitancy |  | - | -.59*** | .52*** | .68*** | -.30*** | .28*** | -.01 |
| 3. General Vaccine Trust |  |  | - | -.33*** | -.48*** | .46*** | -.12*** | .17*** |
| 4. Conspiracy Mentality |  |  |  | - | .59*** | -.17*** | .21*** | -.09*** |
| 5. COVID-19 Conspiracies |  |  |  |  | - | -.23*** | .34*** | -.01 |
| 6. Belief in Science |  |  |  |  |  | - | -.21*** | .09*** |
| 7. Political Orientation |  |  |  |  |  |  | - | .24*** |
| 8. Financial Satisfaction |  |  |  |  |  |  |  | - |

Note. \*\*\* $p < .001$ , \*\* $p < .01$ , \* $p < .05$ ; Political orientation: 1 = left, 10 = right

**Table 3c.** Bivariate correlations between variables (Turkey)

|  | 1 | 2 | 3 | 4 | 5 | 6 | 7 | 8 |
| --- | --- | --- | --- | --- | --- | --- | --- | --- |
| 1. COVID-19 Vaccine Acceptance | - | -.55*** | .54*** | -.21*** | -.23*** | .08** | .12*** | .13*** |
| 2. General Vaccine Hesitancy |  | - | -.53*** | .41*** | .48*** | .06* | -.13*** | -.13*** |
| 3. General Vaccine Trust |  |  | - | -.19*** | -.32*** | .18*** | .03 | .18*** |
| 4. Conspiracy Mentality |  |  |  | - | .36*** | .29*** | -.14*** | -.10*** |
| 5. COVID-19 Conspiracies |  |  |  |  | .- | -.08** | .11*** | -.09*** |
| 6. Belief in Science |  |  |  |  |  | - | -.33*** | .07** |
| 7. Political Orientation |  |  |  |  |  |  | - | .27*** |
| 8. Financial Satisfaction |  |  |  |  |  |  |  | - |

Note. \*\*\* $p < .001$ , \*\* $p < .01$ , \* $p < .05$ ; Political orientation: 1 = left, 10 = right

**Table 4a.** Multiple Regression Models of Vaccine Hesitancy and Vaccine Trust for the UK

|  | General Vaccine Hesitancy |  |  | General Vaccine Trust |  |  |
| --- | --- | --- | --- | --- | --- | --- |
| | $\beta$ | *SE* | *p* | $\beta$ | *SE* | *p* |
| Conspiracy Mentality | .28 | .03 | < .001 | -.07 | .03 | .006 |
| COVID-19 Conspiracy | .46 | .03 | < .001 | -.42 | .03 | < .001 |
| Belief in Science | -.09 | .03 | < .001 | .22 | .03 | < .001 |
| Political Orientation (left to right) | .07 | .01 | < .001 | .08 | .01 | .001 |
| Financial Satisfaction | -.05 | .01 | .007 | .12 | .01 | < .001 |
| Education | .03 | .01 | .196 | -.00 | .01 | .884 |
| Age | .01 | .00 | .628 | .07 | .00 | .004 |
| Gender (0 = Women, 1 = Men) | -.01 | .04 | .614 | -.01 | .05 | .642 |
| Ethnicity (0 = White, 1 = Asian) | .04 | .18 | .023 | .00 | .09 | .997 |
| Ethnicity (0 = White, 1 = Black / African / Caribbean) | .05 | .13 | .005 | -.01 | .13 | .750 |
| Ethnicity (0 = White, 1 = Other) | .01 | .12 | .674 | .00 | .12 | .995 |
| *F* |  | 146.735 |  |  | 79.141 |  |
| *R*<sup>2</sup> |  | .52 |  |  | .36 |  |

**Table 4b.** Multiple Regression Models of Vaccine Hesitancy and Vaccine Trust for the US

|  | General Vaccine Hesitancy |  |  | General Vaccine Trust |  |  |
| --- | --- | --- | --- | --- | --- | --- |
| | $\beta$ | *SE* | *p* | $\beta$ | *SE* | *p* |
| Conspiracy Mentality | .19 | .04 | < .001 | -.03 | .04 | .240 |
| COVID-19 Conspiracy | .52 | .03 | < .001 | -.39 | .03 | < .001 |
| Belief in Science | -.14 | .02 | < .001 | .36 | .03 | < .001 |
| Political Orientation (left to right) | .03 | .01 | .175 | .07 | .01 | .003 |
| Financial Satisfaction | .01 | .01 | .725 | .09 | .01 | < .001 |
| Education | .03 | .02 | .102 | .06 | .02 | .009 |
| Age | -.03 | .00 | .204 | .04 | .00 | .107 |
| Gender (0 = Women, 1 = Men) | .03 | .05 | .132 | .00 | .05 | .960 |
| Ethnicity (0 = White, 1 = Hispanic/Latino) | -.01 | .07 | .669 | .02 | .08 | .329 |
| Ethnicity (0 = White, 1 = Black or African American) | .02 | .07 | .194 | -.01 | .08 | .640 |
| Ethnicity (0 = White, 1 = Asian) | .01 | .10 | .648 | .01 | .11 | .765 |
| Ethnicity (0 = White, 1 = Other) | .00 | .11 | .944 | -.03 | .13 | .189 |
| *F* |  | 131.511 |  |  | 81.493 |  |
| *R*<sup>2</sup> |  | .51 |  |  | .39 |  |

**Table 4c.** Multiple Regression Models of Vaccine Hesitancy and Vaccine Trust for Turkey

|  | General Vaccine Hesitancy |  |  | General Vaccine Trust |  |  |
| --- | --- | --- | --- | --- | --- | --- |
| | $\beta$ | *SE* | *p* | $\beta$ | *SE* | *p* |
| Conspiracy Mentality | .24 | .04 | < .001 | -.14 | .05 | < .001 |
| COVID-19 Conspiracy | .34 | .03 | < .001 | -.22 | .03 | < .001 |
| Belief in Science | -.04 | .03 | .092 | .22 | .04 | < .001 |
| Political Orientation (left to right) | -.14 | .01 | < .001 | .07 | .01 | .005 |
| Financial Satisfaction | -.05 | .01 | .022 | .13 | .01 | < .001 |
| Education | -.04 | .01 | .064 | .08 | .01 | .002 |
| Age | .18 | .00 | < .001 | -.08 | .00 | .005 |
| Gender (0 = Women, 1 = Men) | .07 | .05 | .001 | -.04 | .06 | .066 |
| *F* |  | 104.452 |  |  | 44.196 |  |
| *R*<sup>2</sup> |  | .35 |  |  | .19 |  |

Belief in COVID-19 conspiracies was also the strongest predictor negatively correlated with vaccine trust in all three countries (Figure 1c, Table 4a-c). Although conspiracy mentality was also negatively correlated with vaccine trust, the effect sizes were relatively small and, in the US, non-significant (Table 4a-c).

Both conspiracy mentality and belief in COVID-19 conspiracies had negative effects on the probability of COVID-19 vaccine acceptance (Table 5a-c). Controlling for other factors, the probability of COVID-19 vaccine acceptance decreased by 89%, 127%, and 32% with a one-point increase in agreement with COVID-19 conspiracy statements in the UK, US, and Turkey, respectively. Likewise, a one-point increase in the conspiracy mentality scale decreased the probability of COVID-19 vaccine acceptance by 82%, 25%, and 72% in the UK, US, and Turkey, respectively (Table 5a-c).

**Table 5a.** Logistic regression model for the odds of COVID-19 Vaccine Acceptance in the UK

|  | **Odds ratio**<br>**(CI lower, CI upper)** | **p-value** |
| --- | --- | --- |
| Conspiracy Mentality | 0.55 (0.43, 0.71) | < .001 |
| COVID-19 Conspiracies | 0.53 (0.44, 0.64) | < .001 |
| Belief in Science | 1.76 (1.47, 2.11) | < .001 |
| Political Orientation | 1.00 (0.92, 1.09) | .962 |
| Financial Satisfaction | 1.15 (1.08, 1.23) | < .001 |
| Education | 1.01 (0.91, 1.11) | .873 |
| Age | 1.03 (1.02, 1.04) | < .001 |
| Gender (0 = Women, 1 = Men) | 1.27 (0.91, 1.76) | .159 |
| Ethnicity (0 = White, 1 = Asian) | 0.86 (0.51, 1.44) | .564 |
| Ethnicity (0 = White, 1 = Black) | 0.44 (0.22, 0.91) | .027 |
| Ethnicity (0 = White, 1 = Other) | 1.26 (0.57, 2.77) | .569 |

**Table 5b.** Logistic regression model for the odds of COVID-19 Vaccine Acceptance in the US

|  | **Odds ratio**<br>**(CI lower, CI upper)** | **p-value** |
| --- | --- | --- |
| Conspiracy Mentality | 0.80 (0.65, 0.99) | .039 |
| COVID-19 Conspiracies | 0.44 (0.37, 0.51) | < .001 |
| Belief in Science | 2.19 (1.90, 2.52) | < .001 |
| Political Orientation | 1.03 (0.98, 1.09) | .235 |
| Financial Satisfaction | 1.10 (1.05, 1.16) | < .001 |
| Education | 1.22 (1.12, 1.34) | < .001 |
| Age | 1.02 (1.02, 1.03) | < .001 |
| Gender (0 = Women, 1 = Men) | 1.54 (1.18, 2.01) | .002 |
| Ethnicity (0 = White, 1 = Asian) | 2.00 (1.11, 3.61) | .021 |
| Ethnicity (0 = White, 1 = Black) | 1.12 (0.77, 1.63) | .555 |
| Ethnicity (0 = White, 1 = Hispanic) | 1.93 (1.32, 2.82) | .001 |
| Ethnicity (0 = White, 1 = Other) | 0.72 (0.39, 1.33) | .295 |

**Table 5c.**
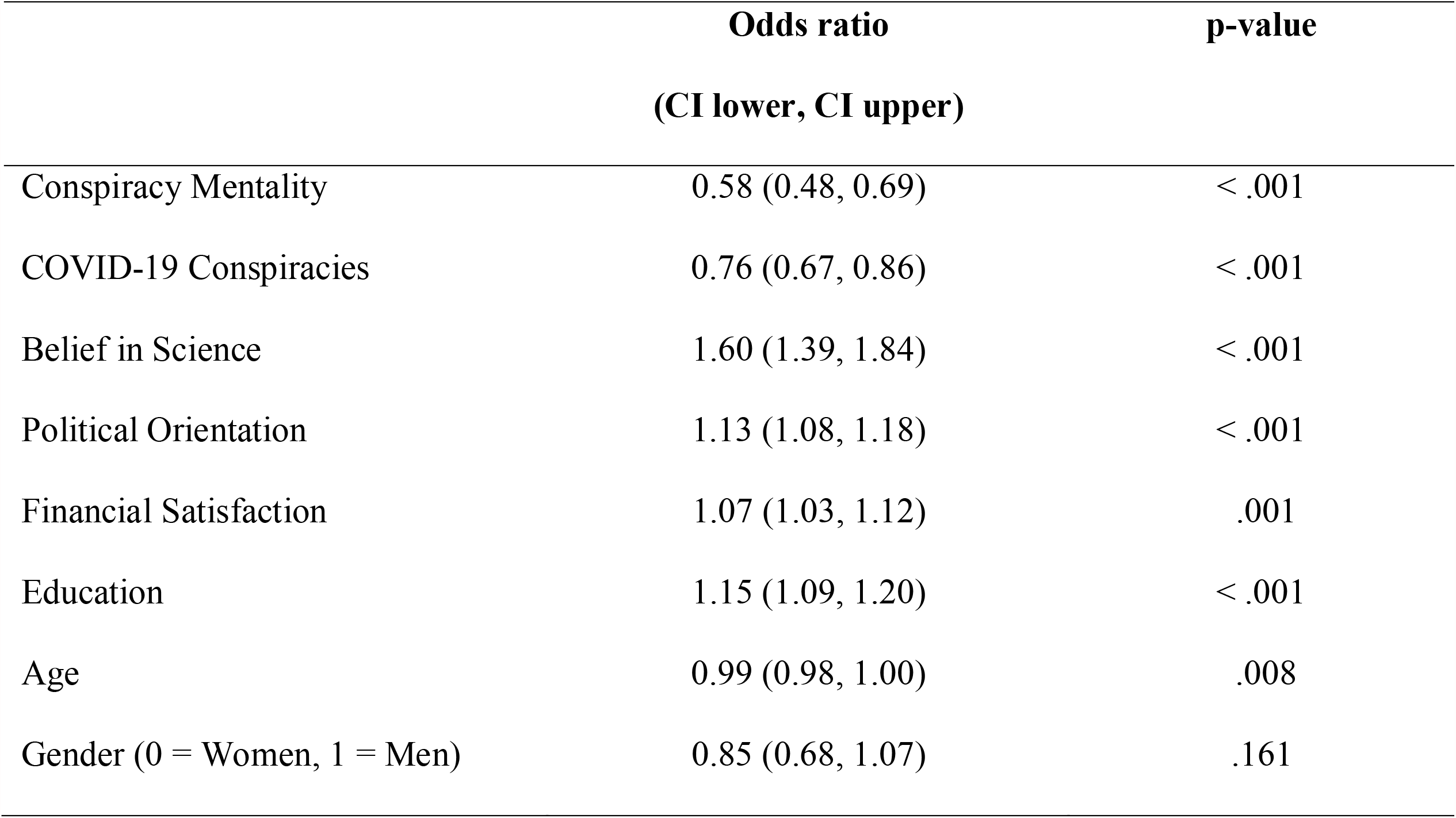
Logistic regression model for the odds of COVID-19 Vaccine Acceptance in Turkey

|  | **Odds ratio**<br>**(CI lower, CI upper)** | **p-value** |
| --- | --- | --- |
| Conspiracy Mentality | 0.58 (0.48, 0.69) | < .001 |
| COVID-19 Conspiracies | 0.76 (0.67, 0.86) | < .001 |
| Belief in Science | 1.60 (1.39, 1.84) | < .001 |
| Political Orientation | 1.13 (1.08, 1.18) | < .001 |
| Financial Satisfaction | 1.07 (1.03, 1.12) | .001 |
| Education | 1.15 (1.09, 1.20) | < .001 |
| Age | 0.99 (0.98, 1.00) | .008 |
| Gender (0 = Women, 1 = Men) | 0.85 (0.68, 1.07) | .161 |

### The level of conspiracy mentality and belief in COVID-19 conspiracies align with the observed cross-cultural variation in vaccine attitudes

There were significant differences in participants’ level of belief in COVID-19 conspiracies across the three countries. The mean agreement score for several COVID-19-related conspiracy statements was highest in Turkey and lowest in the UK (Table 2, Tukey’s HSD: p< 0.001 for UK-US, UK-Turkey and US-Turkey). Participants in the US and Turkey scored higher on the conspiracy mentality scale compared to the participants in the UK (Table 2, Tukey’s HSD: UK-US, p< 0.001, UK-Turkey, p< 0.001, US-Turkey, p= 0.94). The proportion of participants who scored high (4 and above) on the conspiracy mentality and COVID-19 conspiracy belief scales was higher in the US and Turkey compared to the UK (Figure 2).

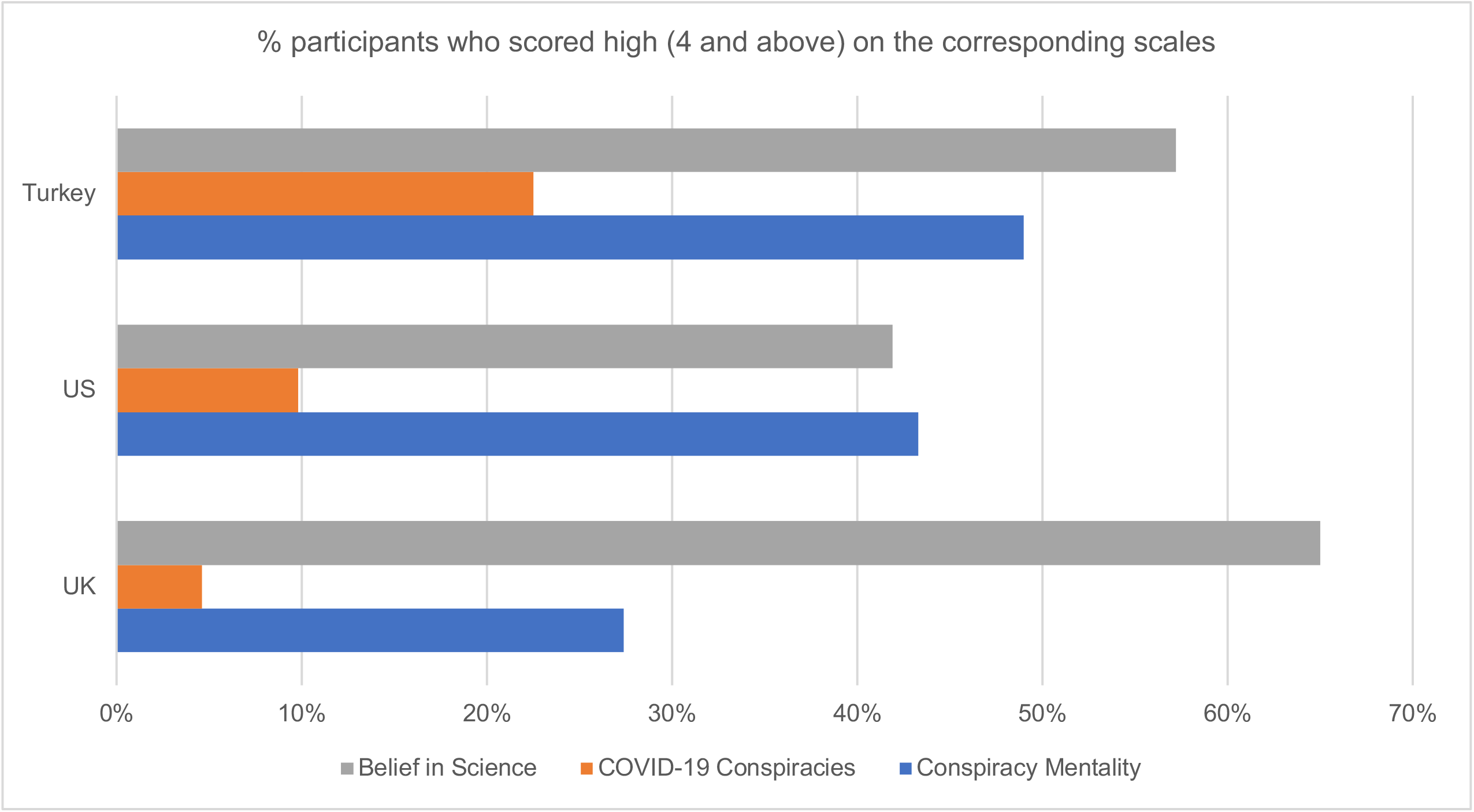

The country-level scores of conspiracy mentality and belief in COVID-19 conspiracies matched with those of vaccination attitudes. For example, participants in the UK scored lowest on agreement with the COVID-19 conspiracy statements and exhibited the highest COVID-19 vaccine acceptance rates, while those in Turkey had the highest COVID-19 conspiracy belief scores and lowest COVID-19 vaccine acceptance.

### Belief in science strongly predicts general vaccine trust and COVID-19 vaccine acceptance, but cannot explain cross-cultural variation

Among the explanatory variables, belief in science exhibited the strongest positive effect on trust in vaccines in all three countries (Figure 1d, Table 4a-c). It also had a strong positive effect on the probability of COVID-19 vaccine acceptance in all three countries (Table 5a-c). A one-point increase in the belief in science scale increased the probability of vaccine acceptance by 76%, 119%, and 60% in the UK, US, and Turkey, respectively (Table 5a-c). Belief in science was negatively correlated with general vaccine hesitancy in the UK and US, but its effect size was smaller compared to those of conspiracy beliefs and conspiracy mentality (Table 4a-b). Belief in science did not predict general vaccine hesitancy in Turkey (Table 4c).

The mean scores for belief in science significantly differed across the UK, US, and Turkey, with participants in the UK scoring highest on this scale (Figure 2, Table 2, Tukey’s HSD: p< 0.001 for UK-US, UK-Turkey). Although the mean levels of vaccine trust and COVID-19 vaccine acceptance were significantly lower in Turkey, participants in Turkey scored significantly higher on belief in science than those in the US (Table 2, Tukey’s HSD: p< 0.001 for US-Turkey), suggesting that belief in science cannot explain the variation in vaccine trust and COVID-19 vaccine acceptance among these countries.

### Demographic variables had a minor effect on vaccine attitudes

Demographic variables had either nonsignificant or small effects on vaccine attitudes compared to conspiracy beliefs and belief in science. The size and direction of the effect differed depending on the country. Below we summarise the predictive power of each control variable on vaccination attitudes across the three countries:

#### Age and gender

The demographic factor that most significantly contributed to vaccine hesitancy in Turkey was age: hesitancy increased with age (Table 4c). Age also had a negative effect on vaccine trust and COVID-19 vaccine acceptance in Turkey (Table 4c-5c). A 10-year increase in age decreased the probability of vaccine acceptance by 12% (Table 5c). In contrast, age did not predict vaccine hesitancy either in the UK or the US (Table 4a-b). Age had a small but positive effect on COVID-19 vaccine acceptance in those two countries (Table 5a-b). Furthermore, gender did not affect vaccine attitudes in the UK (Table 4a-5a). However, in the US, men had a 54% higher probability of COVID-19 vaccine acceptance than women (Table 5b), and in Turkey, general vaccine hesitancy was mildly higher among men than women (Table 4c).

#### Political orientation

In Turkey, political orientation towards the left was positively associated with general vaccine hesitancy, and a one-point increase from left to right political orientation increased the probability of COVID-19 vaccine acceptance by 13% (Table 4c-5c). Political orientation did not affect the probability of vaccine acceptance either in the UK or the US (Table 5a-b). It had a small effect on vaccine trust in all three countries (Table 4a-c). To further understand this effect, we divided participants into three groups on the political orientation scale: left (0-3), centre (4-6), and right (7-10). We then calculated the mean level of trust in vaccines for each group. The mean levels of vaccine trust for left, centre, and right-wing political views were 4.9, 4.6, and 4.8 for participants in the UK; 4.7, 4.3, and 4.2 for those in the US; and 4.3, 4.1, and 4.3 for those in Turkey. Therefore, participants only slightly differed in vaccine trust based on their political views, and there was no clear pattern, with the exception of the US.

#### Level of education and financial satisfaction

Level of education was not correlated with general vaccine hesitancy in any of the countries (Table 4a-c). It had a positive effect on vaccine trust and COVID-19 vaccine acceptance in Turkey and the US, but not in the UK (Tables 4 and 5). Vaccine trust increased with increasing financial satisfaction in all countries (Table 4a-c). Financial satisfaction was negatively correlated with vaccine hesitancy in the UK and Turkey, with small effects, but not in the US (Table 4a-c). Financial satisfaction increased the probability of COVID-19 vaccine acceptance in all three countries (Table 5a-c).

#### Ethnicity

Vaccine hesitancy among Black/African/Caribbean and Asian British people was higher compared to White British people; however, the effect sizes were small (Table 4a). The probability of COVID-19 vaccine acceptance was lower among Black/African/Caribbean British participants (Table 5a). Ethnicity did not affect general vaccine trust in the UK (Table 4a). It affected neither general vaccine hesitancy nor trust in the US (Table 4b). The probability of COVID-19 vaccine acceptance increased if a person had Asian or Hispanic ethnic origin in the US (Table 5b).

## Discussion

In this study, we examined the variation in vaccine attitudes in the UK, US, and Turkey. Trust in vaccines and COVID-19 vaccine acceptance were both highest in the UK and lowest in Turkey. While general vaccine hesitancy was lowest in the UK, it did not differ between Turkey and the US. In line with our predictions, conspiratorial thinking—especially the level of agreement with COVID-19 conspiracy statements—was strongly associated with vaccine hesitancy and its variation across the three countries. Belief in science was positively associated with trust in vaccines and increased the probability of COVID-19 vaccine acceptance; however, it did not explain the observed differences in vaccine attitudes across the three countries.

We hypothesised that different measures of vaccine attitude might be influenced by different factors. Our findings indicate that while belief in science is a strong predictor of trust in the safety and effectiveness of vaccines and the acceptance of new vaccines, it is not as strongly correlated with general vaccine hesitancy (which in our study captured the degree of concern around side effects, pharmaceutical financial profits, and preference for natural immunity). Instead, variation in vaccine hesitancy, vaccine trust, and acceptance of new vaccines in the UK, US, and Turkey strongly aligned with the variation in the level of conspiracy mentality and belief in COVID-19 conspiracies in those countries. Therefore, it is likely that the low vaccine acceptance rates in certain regions, like the Middle East, are driven by high levels of conspiratorial thinking (Sallam et al., 2021; Zonis & Joseph, 1994). Moreover, although a recent study found that a societal consensus on trust in science reflects country-level vaccine confidence (Sturgis et al., 2021), our findings suggest that positive perceptions of science alone cannot explain the cross-cultural variation in vaccine attitudes. One reason may be that people’s belief in science (how much they think science can help humans attain knowledge or understand the universe) may not always reflect their scientific literacy or analytical thinking. If a person believes in the ability of science to attain truth but is inclined to intuitive thinking as opposed to analytical thinking, he or she will still be more susceptible to conspiracy beliefs (Alper, Bayrak, & Yilmaz, 2020; Swami, Voracek, Stieger, Tran, & Furnham, 2014). In fact, this theory may explain why people in Turkey scored higher on belief in science compared to people in the US but still exhibited the lowest level of trust in vaccines. Moreover, a recent study showed that trust in science makes people more vulnerable to pseudoscience and increases their chance of believing false claims that contain scientific references (O’Brien, Palmer, & Albarracin, 2021). Together these findings suggest that the negative effect of misinformation and conspiracy beliefs overrides the positive effect of belief in science on vaccine trust.

Although a general conspiracy mentality and belief in COVID-19 conspiracies were highly correlated, conspiracy beliefs specific to COVID-19 had a stronger correlation with vaccine hesitancy and general trust in vaccines. Moreover, in Turkey, conspiracy mentality and belief in science were positively correlated, while COVID-19 conspiracy beliefs were negatively correlated with belief in science (Table 3a-c). In all three countries, the correlation between conspiracy mentality and COVID-19 conspiracy beliefs were positive and significant; however, the correlations were stronger in the UK and US compared to Turkey (Table 3a-c). In Turkey, conspiracy mentality was positively associated with left-wing political orientation; in the US, conspiracy mentality was positively associated with right-wing orientation. There was no association in the UK (Table 3a-c). Moreover, there was a positive correlation between right-wing political orientation and endorsement of COVID-19 conspiracies in all three countries. These findings suggest that the aetiology of conspiracy mentality and belief in specific COVID-19 conspiracy statements may be different, especially in Turkey, and warrants further investigation.

The relatively small effects of several demographic factors in our study reinforce previous findings indicating that belief in conspiracies is the strongest predictor of vaccination attitudes across cultures and that demographic variables have trivial effects (Hornsey et al., 2018). Moreover, the direction and magnitude of the effect of the demographic variables differed between countries. For example, while older participants were more hesitant about vaccines and less willing to get vaccinated against COVID-19 in Turkey than younger participants, this was not the case in the UK or the US. Moreover, political orientation exhibited contrasting effects on vaccine hesitancy in Turkey: left-wing people were found to be more vaccine hesitant on average than right-wing people. We suspect this result was driven by the significant correlation between conspiracy mentality and political orientation in the country (Table 3c). These findings suggest that research and policies should focus on understanding the psychological roots of vaccine attitudes, more so than demographic factors, to tackle vaccine hesitancy globally. Studies exploring the psychological correlates of conspiracy beliefs are especially crucial, as conspiracy beliefs can be a mediator between those correlates and vaccine attitudes.

It is important to note that at the time of our study (March-April 2021), the percentage of participants who had already received a COVID-19 vaccine was low in Turkey (1.4%) compared to the UK (24.3%) and the US (14.8%), largely due to differences in vaccine supply. It is possible that some of the vaccine-hesitant people in Turkey will decide to get vaccinated against COVID-19 as more vaccines become available and more people receive them. However, some may also decide not to get vaccinated, especially given that undecided individuals are more likely to come across anti-vaccination clusters in online networks (Johnson et al., 2020). How vaccination decisions change when vaccine programs are ongoing is an intriguing question. It is striking that the percentage of participants in the UK who were willing to get vaccinated against COVID-19 (83%) in our previous survey in May 2020, when vaccines were not yet available, remained the same in this study (the combined percentage of people who already got vaccinated and those who were willing to be vaccinated was 83% in this study; Salali & Uysal, 2020). Nevertheless, the vaccination programs may halt, as the remaining unvaccinated people will contain a high percentage of people who refuse to get vaccinated. It is alarming that 18.3% participants in the US (as opposed to 11.6% in Turkey and 6.1% in the UK) indicated that they are not willing to get vaccinated against COVID-19.

If conspiracy beliefs are the strongest predictor of vaccine hesitancy, how can we tackle them? Trying to reduce people’s belief in conspiracies through corrective information is often, if not always, fruitless (Hornsey, 2020; Hornsey et al., 2018). Moreover, people tend to cherry-pick information that supports their existing views and reject the information that challenges them (motivated reasoning; (Kunda, 1990), which makes approaches such as fact-checking ineffective (Hornsey, 2020). Given that the origin of the novel coronavirus is still uncertain (Mallapaty, 2021), other solutions than challenging conspiracy beliefs may be more effective for tackling COVID-19 vaccine hesitancy. For example, encouraging people to become vaccinated by making vaccinations free and easy to access is one potential strategy (Chevallier, Hacquin, & Mercier, 2021). Nudges, such as text messages, to remind people that their shot is reserved for them have been found to increase flu jab intake and may also be effective at encouraging “mildly” hesitant people (Milkman et al., 2021).

Moreover, “good of the group” arguments may not prove successful if the perceived long-term costs to the individual are high and if the argument is not coming from an in-group member (Arnot et al., 2020). For example, a person worried about a new vaccine’s long-term side effects may not care much about vaccine promotion messages encouraging people to help protect others. Individual and family-based incentives may be more effective (Jarrett et al., 2015). Although controversial, mandatory vaccine certificates for travelling may also encourage vaccine-hesitant people, but not those who actively refuse vaccinations, to get vaccinated (Osama, Razai, & Majeed, 2021). In addition, not only the content of the vaccination message, but where it comes from is important. People trust in and conform to behaviours of those with shared identities; therefore, vaccine information coming from an in-group member may be more effective than information coming from an out-group member, especially when tackling hesitancy in minority groups (Arnot et al., 2020). Likewise, encouraging early adopters of new vaccines to announce their pro-vaccine intention may help to build a social norm of vaccination, as people learn about and conform to norms by observing their peers (Arnot et al., 2020; Chevallier et al., 2021).

In this paper we demonstrated that belief in conspiracies is the strongest predictor of vaccine hesitancy and explains the variation in vaccine attitudes across countries. Studies exploring the psychological roots of those beliefs are therefore crucial to achieve a deep understanding of vaccine attitudes.

## Data Availability

All data files and R codes are available at Open Science Framework.

https://osf.io/hcvny/

## Author contributions

GDS conceived the project. All authors contributed to the survey design and statistical analyses. GDS wrote the paper with the help of MSU.

## Data availability

All data files and R codes are available at https://osf.io/hcvny/.

## References

Alper, S., Bayrak, F., & Yilmaz, O. (2020). Psychological correlates of COVID-19 conspiracy beliefs and preventive measures: Evidence from Turkey. Current Psychology. https://doi.org/10.1007/s12144-020-00903-0

Anderson, R. M., Vegvari, C., Truscott, J., & Collyer, B. S. (2020). Challenges in creating herd immunity to SARS-CoV-2 infection by mass vaccination. The Lancet, 396(10263), 1614–1616. https://doi.org/10.1016/S0140-6736(20)32318-7

Arnot, M., Brandl, E., Campbell, O. L. K., Chen, Y., Du, J., Dyble, M., … Zhang, H. (2020). How evolutionary behavioural sciences can help us understand behaviour in a pandemic. Evolution, Medicine and Public Health, 2020(1), 264–278. https://doi.org/10.1093/EMPH/EOAA038

Bertin, P., Nera, K., & Delouvée, S. (2020). Conspiracy Beliefs, Rejection of Vaccination, and Support for hydroxychloroquine: A Conceptual Replication-Extension in the COVID-19 Pandemic Context. Frontiers in Psychology, 11, 565128. https://doi.org/10.3389/fpsyg.2020.565128

Bono, S. A., Faria de Moura Villela, E., Siau, C. S., Chen, W. S., Pengpid, S., Hasan, M. T., … Colebunders, R. (2021). Factors Affecting COVID-19 Vaccine Acceptance: An International Survey among Low-and Middle-Income Countries. Vaccines, 9(5), 515. https://doi.org/10.3390/vaccines9050515

Bruder, M., Haffke, P., Neave, N., Nouripanah, N., & Imhoff, R. (2013). Measuring Individual Differences in Generic Beliefs in Conspiracy Theories Across Cultures: Conspiracy Mentality Questionnaire. Frontiers in Psychology, 4, 225. https://doi.org/10.3389/fpsyg.2013.00225

Chevallier, C., Hacquin, A. S., & Mercier, H. (2021). COVID-19 Vaccine Hesitancy: Shortening the Last Mile. Trends in Cognitive Sciences, 25(5), 331–333. https://doi.org/10.1016/j.tics.2021.02.002

de Figueiredo, A., Simas, C., Karafillakis, E., Paterson, P., & Larson, H. J. (2020). Mapping global trends in vaccine confidence and investigating barriers to vaccine uptake: a large-scale retrospective temporal modelling study. The Lancet, 396(10255), 898–908. https://doi.org/10.1016/S0140-6736(20)31558-0

Farias, M., Newheiser, A. K., Kahane, G., & de Toledo, Z. (2013). Scientific faith: Belief in science increases in the face of stress and existential anxiety. Journal of Experimental Social Psychology, 49(6), 1210–1213. https://doi.org/10.1016/j.jesp.2013.05.008

Freeman, D., Loe, B. S., Chadwick, A., Vaccari, C., Waite, F., Rosebrock, L., … Lambe, S. (2021). COVID-19 vaccine hesitancy in the UK: The Oxford coronavirus explanations, attitudes, and narratives survey (Oceans) II. Psychological Medicine, 1–15. https://doi.org/10.1017/S0033291720005188

Freeman, D., Waite, F., Rosebrock, L., Petit, A., Causier, C., East, A., … Lambe, S. (2020). Coronavirus Conspiracy Beliefs, Mistrust, and Compliance with Government Guidelines in England. Psychological Medicine, 1–13. https://doi.org/10.1017/S0033291720001890

Gallup. (2019). Wellcome Global Monitor – First Wave Findings.

Goertzel, T. (1994). Belief in conspiracy theories. Political Psychology, (15), 731–742.

Hornsey, M. J. (2020). Why Facts Are Not Enough: Understanding and Managing the Motivated Rejection of Science. Current Directions in Psychological Science, 29(6), 583–591. https://doi.org/10.1177/0963721420969364

Hornsey, M. J., Harris, E. A., & Fielding, K. S. (2018). The psychological roots of antivaccination attitudes: A 24-nation investigation. Health Psychology, 37(4), 307–315. https://doi.org/10.1037/hea0000586

Jarrett, C., Wilson, R., O’Leary, M., Eckersberger, E., Larson, H. J., Eskola, J., … Schuster, M. (2015). Strategies for addressing vaccine hesitancy - A systematic review. Vaccine, 33(34), 4180–4190. https://doi.org/10.1016/j.vaccine.2015.04.040

Johnson, N. F., Velásquez, N., Restrepo, N. J., Leahy, R., Gabriel, N., El Oud, S., … Lupu, Y. (2020). The online competition between pro-and anti-vaccination views. Nature, 582(7811), 230–233. https://doi.org/10.1038/s41586-020-2281-1

Jolley, D., & Douglas, K. M. (2014). The effects of anti-vaccine conspiracy theories on vaccination intentions. PLoS ONE, 9(2). https://doi.org/10.1371/journal.pone.0089177

Kunda, Z. (1990). The case for Motivated Reasoning. Psychological Bulletin, 108(3), 480– 498.

Kwok, K. O., Lai, F., Wei, W. I., Wong, S. Y. S., & Tang, J. W. T. (2020). Herd immunity – estimating the level required to halt the COVID-19 epidemics in affected countries. Journal of Infection. https://doi.org/10.1016/j.jinf.2020.03.027

Lazarus, J. V., Ratzan, S. C., Palayew, A., Gostin, L. O., Larson, H. J., Rabin, K., … El-Mohandes, A. (2021). A global survey of potential acceptance of a COVID-19 vaccine. Nature Medicine, 27(2), 225–228. https://doi.org/10.1038/s41591-020-1124-9

Lewandowsky, S., Gignac, G. E., & Oberauer, K. (2013). The Role of Conspiracist Ideation and Worldviews in Predicting Rejection of Science. PLoS ONE, 8(10). https://doi.org/10.1371/journal.pone.0075637

Mallapaty, S. (2021). What’s next in the search for COVID’s origins. Nature, 592, 337–338.

Martin, L. R., & Petrie, K. J. (2017). Understanding the Dimensions of Anti-Vaccination Attitudes: the Vaccination Attitudes Examination (VAX) Scale. Annals of Behavioral Medicine, 51(5), 652–660. https://doi.org/10.1007/s12160-017-9888-y

Milkman, K. L., Patel, M. S., Gandhi, L., Graci, H. N., Gromet, D. M., Ho, H., … Duckworth, A. L. (2021). A megastudy of text-based nudges encouraging patients to get vaccinated at an upcoming doctor’s appointment. Proceedings of the National Academy of Sciences of the United States of America, 118(20), 10–12. https://doi.org/10.1073/PNAS.2101165118

O’Brien, T. C., Palmer, R. P., & Albarracin, D. (2021). Misplaced Trust: When Trust in Science Fosters Belief in Pseudoscience and the Benefits of Critical Evaluation. Journal of Experimental Social Psychology, 96, 104184. https://doi.org/10.1016/j.jesp.2021.104184

Osama, T., Razai, M. S., & Majeed, A. (2021). Covid-19 vaccine passports: Access, equity, and ethics. The BMJ, 373, 1–2. https://doi.org/10.1136/bmj.n861

Roozenbeek, J., Schneider, C. R., Dryhurst, S., Kerr, J., Freeman, A. L. J., Recchia, G., … Van Der Linden, S. (2020). Susceptibility to misinformation about COVID-19 around the world: Susceptibility to COVID misinformation. Royal Society Open Science, 7(10). https://doi.org/10.1098/rsos.201199

Salali, G. D., & Uysal, M. S. (2020). COVID-19 vaccine hesitancy is associated with beliefs on the origin of the novel coronavirus in the UK and Turkey. Psychological Medicine, 1– 3.

Sallam, M. (2021). Covid-19 vaccine hesitancy worldwide: A concise systematic review of vaccine acceptance rates. Vaccines, 9(2), 1–15. https://doi.org/10.3390/vaccines9020160

Sallam, M., Dababseh, D., Eid, H., Al-Mahzoum, K., Al-Haidar, A., Taim, D., … Mahafzah, A. (2021). High rates of covid-19 vaccine hesitancy and its association with conspiracy beliefs: A study in jordan and kuwait among other arab countries. Vaccines, 9(1), 1–16. https://doi.org/10.3390/vaccines9010042

Simas, C., & Larson, H. J. (2021). Overcoming vaccine hesitancy in low-income and middleincome regions. Nature Reviews Disease Primers, 7(1), 1–2. https://doi.org/10.1038/s41572-021-00279-w

Sturgis, P., Brunton-Smith, I., & Jackson, J. (2021). Trust in science, social consensus and vaccine confidence. Nature Human Behaviour. https://doi.org/10.1038/s41562-021-01115-7

Swami, V., Voracek, M., Stieger, S., Tran, U. S., & Furnham, A. (2014). Analytic thinking reduces belief in conspiracy theories. Cognition, 133(3), 572–585. https://doi.org/10.1016/j.cognition.2014.08.006

Zonis, M., & Joseph, C. M. (1994). Conspiracy Thinking in the Middle East. Political Psychology, 15(3), 443–459.

